# Hypertensive adults exhibit lower myelin content: A multicomponent relaxometry and diffusion MRI study

**DOI:** 10.1101/2023.02.21.23286279

**Authors:** John P. Laporte, Mary E. Faulkner, Zhaoyuan Gong, Mohammad A.B.S. Akhonda, Luigi Ferrucci, Josephine M. Egan, Mustapha Bouhrara

## Abstract

It is unknown whether hypertension plays any role in cerebral myelination. To fill this knowledge gap, we studied ninety cognitively unimpaired adults, age range 40 to 94 years, that are participants in the Baltimore Longitudinal Study of Aging (BLSA) and the Genetic and Epigenetic Signatures of Translational Aging Laboratory Testing (GESTALT) to look for potential associations between hypertension and cerebral myelin content across fourteen white matter brain regions. Myelin content was probed using our advanced multicomponent magnetic resonance (MR) relaxometry method of myelin water fraction (MWF), a direct and specific MR imaging (MRI) measure of myelin content, and longitudinal and transverse relaxation rates (*R*_*1*_ and *R*_*2*_), two highly sensitive MRI metrics of myelin content. We also applied diffusion tensor imaging (DTI) MRI to measure fractional anisotropy (FA), mean diffusivity (MD), radial diffusivity (RD) and axial diffusivity (AxD) values, which are metrics of cerebral microstructural tissue integrity, to provide contact with previous MRI findings. After adjustment of age, sex, systolic blood pressure, smoking status, diabetes status and cholesterol level, our results indicated that participants with hypertension exhibited lower MWF, FA, *R*_*1*_ and *R*_*2*_ values and higher MD, RD and AxD values, indicating lower myelin content and higher impairment to the brain microstructure. These associations were significant across several white matter regions, particularly in the corpus callosum, fronto-occipital fasciculus, temporal lobes, internal capsules, and corona radiata. These original findings suggest a direct association between myelin content and hypertension, and form the basis for further investigations including longitudinal assessments of this relationship.

## 1. INTRODUCTION

Hypertension is the primary risk factor for stroke, ischemic white matter lesions, infarcts, and atherosclerosis, as well as cardiovascular and microvascular diseases (1-3). Further, hypertension is the most widely accepted risk factor associated with a myriad of neurodegenerative diseases, especially Alzheimer’s disease and related dementias (4). Emerging evidence suggests that hypertension leads to vessel wall remodeling, potentially leading to hypoperfusion and associated hypoxia, as well as reduced glucose transport into the brain with concomitant accelerated cerebral tissue degeneration (5, 6). Indeed, this paradigm is further supported by recent longitudinal studies revealing a direct association between hypertension in midlife and reduced cerebral blood flow in later life (7, 8). Therefore, examining the extent of any possible association between hypertension and cerebral tissue integrity is paramount for our global understanding of neurodegenerative disease risk factors and progression.

In recent years, magnetic resonance imaging (MRI) studies, based extensively on diffusion tensor imaging (DTI), have revealed an association between hypertension and abnormal cerebral microstructural white matter integrity (8). DTI is an MRI technique sensitive to the underlying microarchitecture of the brain parenchyma and the degree and direction of water molecule mobility. These studies have documented that hypertension, indicated by a blood pressure >140/90 mmHg or the use of anti-hypertensive medication, is associated with lower values of fractional anisotropy (FA) and higher values of mean diffusivity (MD), radial diffusivity (RD) and axial diffusivity (AxD) (9-11). Reduced FA concomitant with an increase in RD is associated with demyelination (12), whereas reduced FA in conjunction with increased AxD is believed to be associated with axonal injury or death (13). These changes in cerebral microstructural integrity associated with hypertension have been interpreted as deterioration in axonal myelination. However, although DTI metrics such as FA and MD are sensitive to cerebral microstructural changes, they are not specific. Indeed, there are multiple methodological and biological factors that can affect the DTI-derived eigenvalues from which the DTI indices are calculated (14, 15); these include, but not limited to, axonal degeneration, flow, temperature, hydration, macromolecular content and architectural features, including fiber fanning or crossing. Therefore, to our knowledge, the association between hypertension and myelination has not yet been established. To address this limitation, multicomponent relaxometry methods provide a greater specificity to quantify myelin content in white matter and probe related changes that occur during brain development and neurodegenerative diseases (16, 17). Multicomponent relaxometry separates the measured MR signal in white matter into two pools of water, namely the intra- and extracellular water pool and the water trapped between the myelin sheaths calculated as the myelin water fraction (MWF) (18, 19). MWF is an *in vivo* specific index of myelin content and is a potential marker for myelin alterations. To the best of our knowledge, no MR studies have employed multicomponent relaxometry analysis, specifically MWF imaging, to investigate the relationship between myelin content and hypertension in aging adults.

In this study, we examined the association between hypertension and myelin content as probed using MWF on a cohort of well-characterized cognitively unimpaired adults (*N* = 90), across the age range of 40 to 94 years. Each participant underwent our Bayesian Monte Carlo (BMC)-mcDESPOT protocol for MWF as a direct measure of myelin content, as well as mapping of longitudinal and transverse relaxation rates (*R*_*1*_ and *R*_*2*_) as sensitive but non-specific measures of myelin content (6, 20, 21). Indeed, *R*_*1*_ and *R*_*2*_ values depend on both water mobility and macromolecular tissue composition, including local lipid and iron content, the main constituents of myelin, and thus are expected to be directly associated with differences in myelin content. To establish a connection with previous MRI findings, participants have undergone our additional DTI protocol for FA, MD, RD and AxD mapping (22).

## 2. MATERIAL & METHODS

### 2.1. Study Cohort

Participants are volunteers of the Baltimore Longitudinal Study of Aging (BLSA) and the Genetic and Epigenetic Signatures of Translational Aging Laboratory Testing (GESTALT) studies (23, 24). Both BLSA and GESTALT seek to evaluate multiple biomarkers associated with aging, with essentially identical inclusion and exclusion criteria. Participants with metallic implants or major neurologic or medical disorders are excluded on first admission. All participants were administered the Mini Mental State Examination (MMSE). Informed consent was obtained from participants prior to the conduct of the experiments, in compliance with the local Institutional Review Board.

### 2.2. Data Acquisition

MRI scans were performed on a 3T whole body Philips MRI system (Achieva, Best, The Netherlands) using the internal quadrature body coil for transmission and an eight-channel phased-array head coil for reception. Each participant underwent our BMC-mcDESPOT protocol for MWF, *R*_*1*_, and *R*_*2*_ mapping (16, 25). This imaging protocol consisted of 3D spoiled gradient recalled echo (SPGR) images acquired with flip angles (FAs) of [2 4 6 8 10 12 14 16 18 20]°, echo time (TE) of 1.37 ms, repetition time (TR) of 5 ms and acquisition time of ∼5 min, as well as 3D balanced steady-state free precession (bSSFP) images acquired with FAs of [2 4 7 11 16 24 32 40 50 60]°, TE of 2.8 ms, TR of 5.8 ms, and acquisition time of ∼6 min. The bSSFP images were acquired with radiofrequency (RF) excitation pulse phase increments of 0 or 180° to account for off-resonance effects, with a total scan time of ∼12 min (∼6 min for each phase-cycling scan). All SPGR and bSSFP images were acquired with an acquisition matrix of 150 × 130 × 94, voxel size 1.6 mm × 1.6 mm × 1.6 mm. To correct for excitation RF inhomogeneity (26, 27), we used the double-angle method (DAM), which consisted of acquiring two fast spin-echo images with FAs of 45° and 90°, TE of 102 ms, TR of 3000 ms, acquisition voxel size of 2.6 mm × 2.6 mm × 4 mm, and acquisition time of ∼4 min. The total acquisition time was ∼21 min. All images were acquired with field-of-view of 240 mm × 208 mm × 150 mm, SENSE factor of 2, and reconstructed to a voxel size of 1 mm × 1 mm × 1 mm. We emphasize that all MRI studies and ancillary measurements were performed with the same MRI system, with the same pulse sequences, and at the same facility for both BLSA and GESTALT participants.

The DTI protocol consisted of diffusion-weighted images (DWI) acquired with single-shot EPI, TR of 10 s, TE of 70 ms, two *b*-values of 0 and 700 s/mm^2^, with the latter encoded in 32 directions, acquisition matrix of 120 × 104 × 75, and acquisition voxel size of 2 mm × 2 mm × 2 mm. All images were acquired with field of view of 240 mm × 208 mm × 150 mm.

### 2.3. Data Processing

For each participant, using the FLIRT analysis as implemented in the FSL software (28), all SPGR, bSSFP, or DAM images were linearly registered to the SPGR image obtained at FA of 8° and the respective derived transformation matrices were then applied to the original SPGR, bSSFP, or DAM images. Then, a whole-brain MWF map was generated using BMC-mcDESPOT from these co-registered SPGR, bSSFP, and DAM datasets (6, 16, 20). BMC-mcDESPOT assumes a two-relaxation time components system consisting of a short component, attributed to myelin water, and a long component, attributed to intra- and extracellular water. We used the signal model explicitly accounting for non-zero TE (6, 16, 20). This emerging method offers rapid and reliable whole brain MWF map within feasible clinical time (6, 16, 20, 29-31), and has been used in assessing myelin loss in mild cognitive impairment and dementias, as well as examining factors influencing cerebral myelination in normative aging (16, 22, 25, 32-36). A whole-brain *R*_*1*_ map was also generated from the co-registered SPGR and DAM datasets using DESPOT1 (21), and a whole-brain *R*_*2*_ map was generated from the co-registered bSSFP and DAM datasets using DESPOT2 (21). The DW images were corrected for eddy current and motion effects using the affine registration tools as implemented in FSL (28) and registered to the DW image obtained with *b* = 0 s/mm^2^ using FNIRT. We used the DTIfit tool implemented in FSL to calculate the eigenvalue maps which were used to calculate FA, RD, MD and AxD (37).

Further, using FSL software (28), the averaged SPGR image over FAs underwent nonlinear registration to the Montreal Neurological Institute (MNI) standard space, and the computed transformation matrix was then applied to the corresponding DTI indices, MWF, *R*_*1*_, and *R*_*2*_ maps. Fourteen white matter (WM) regions of interest (ROIs) were defined from the MNI structural atlas corresponding to the whole brain (WB), the frontal (FL), parietal (PL), temporal (TL), and occipital (OL) lobes, cerebellum (CRB), corpus callosum (CC), internal capsule (IC), cerebral peduncle (CP), corona radiata (CR), thalamic radiation (TR), fronto-occipital fasciculus (FOF), longitudinal fasciculus (LF), and forceps (F). ROIs were eroded to reduce partial volume effect. Within each ROI, the mean FA, RD, MD, AxD, MWF, *R*_*1*_, and *R*_*2*_ values were calculated.

### 2.4. Systolic and Diastolic blood pressure

Systolic and diastolic blood pressures were recorded three times in both arms in a seated position using a mercury sphygmomanometer sized to the arm of each participant, and the mean of the systolic and diastolic measurements were used in the subsequent analyses (38). Hypertension was defined as a systolic blood pressure greater than or equal to 140 mmHg, a diastolic blood pressure greater than or equal to 90 mmHg, or the use of prescription hypertension medications.

### 2.5. Statistical analysis

To investigate the effect of hypertension on relaxometry (MWF, *R*_*1*_, *R*_*2*_) and diffusion (FA, MD, RD, AxD) MRI metrics, a multiple linear regression analysis was applied using MWF, *R*_*1*_, *R*_*2*_, FA, MD, RD, or AxD within each ROI as the dependent variable and hypertension status, smoking status, systolic blood pressure (SBP), diabetes, cholesterol, age, and sex as independent variables. In all cases, the threshold for statistical significance was *p* < 0.05, while for close-to-significance was taken as *p* < 0.1 after correction for multiple ROI comparisons using the false discovery rate (FDR) method (39, 40). All calculations were performed with MATLAB software (MathWorks, Natick, MA, USA).

## 3. RESULTS

### 3.1. Participants demographic characteristics

Demographic characteristics of the participants are shown in Table 1. After restricting the age range to participants of 40+ years and excluding 6 participants with either cognitive impairment, missing data or bad quality images due to severe motion artifacts, the final cohort consisted of 90 cognitively unimpaired volunteers (mean ± standard deviation MMSE = 28.8 ± 1.3) ranging in age from 40 to 94 years (64.6 ± 17.1 years). Of this cohort, 49 (54.4%) were men, 30 (33.3%) were identified as cigarette smokers while 59 (65.6%) as nonsmokers. Among the cohort, 27 were hypertensive (30.0%), 23 of which taking antihypertensive medication. This cohort also included 3 participants (3.3%) with diabetes (who were also hypertensive), while 87 participants were nondiabetic (96.7%). The mean ± standard deviation values of the systolic blood pressure (SBP) and diastolic blood pressure (DBP) were 117.6 ± 14.5 and 68.5 ± 8.6, respectively. Finally, mean ± standard deviation values of cholesterol level were 183.0 ± 35.1.

**Table 1.** Demographic characteristics of participants of the study cohort.

|  |  |
| --- | --- |
| **Total Sample** | *N* = 90 |
| **Age (yrs.), mean ± SD (*min-max*)** | 64.6 ± 17.1 (40-94) |
| **Sex** |  |
| Male, *N*(%) | 49 (54.4%) |
| Female, *N*(%) | 41 (45.6%) |
| **Smoker** |  |
| Smokers, *N*(%) | 30 (33.3%) |
| Non-smokers, *N*(%) | 59 (65.6%) |
| Other, *N*(%) | 1 (1.1%) |
| **Diabetes** |  |
| Diabetic, *N*(%) | 3(3.3%) |
| Nondiabetic, *N*(%) | 87 (96.7%) |
| **Hypertensive** |  |
| Hypertensive, *N*(%) | 27 (30.0%) |
| Non-hypertensive, *N*(%) | 63 (70.0 %) |
| **SBP (mmHg), mean ± SD (*min-max*)** | 117.6 ± 14.5 (89-161) |
| **DBP (mmHg), mean ± SD (*min-max*)** | 68.5 ± 8.6 (50-87) |
| **Cholesterol, mean ± SD (*min-max*)** | 183.0 ± 35.1 (116-270) |

*SBP*, systolic blood pressure; *DBP*, diastolic blood pressure; *SD*, standard deviation; *min*, minimum; *max*, maximum.

### 3.2 Associations between hypertension and cerebral microstructure

Figure 1 shows the MWF, *R*_*1*_ and *R*_*2*_ relaxometry parameter maps of either hypertensive or non-hypertensive participants within an age range of 70-94 years. This limited age range minimizes the potential effect of age on derived MR parameter maps for this qualitative analysis (statistical quantification of the effect of age as a covariate will be presented below (Table 2 and 3)). Visual inspection indicates that, overall, hypertensive participants exhibit lower regional MWF, *R*_*1*_ and *R*_*2*_ values as compared to non-hypertensive participants. These qualitative results suggest a potentially strong association between hypertension and myelin content.

**Table 2.**
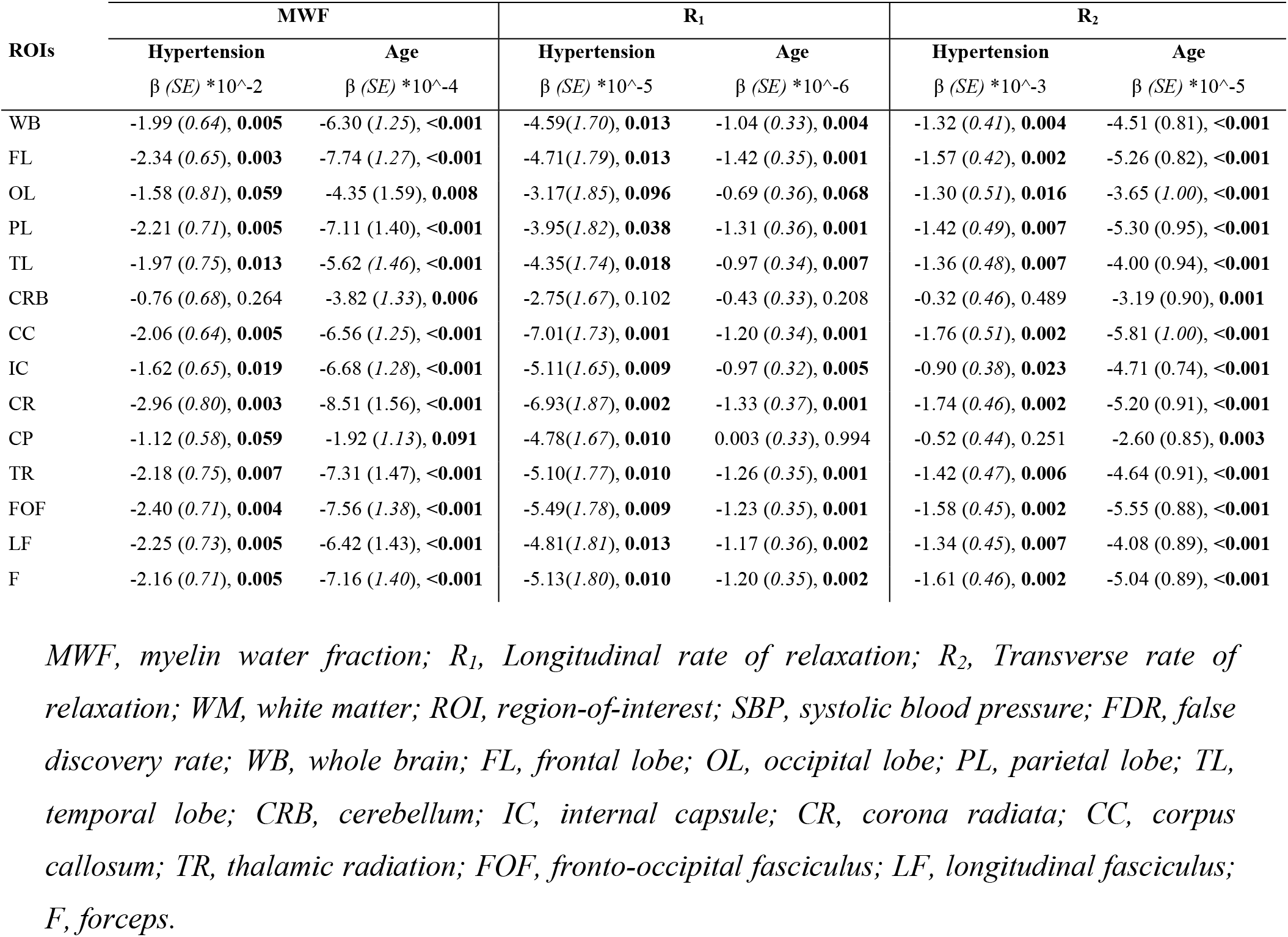
Regression coefficient (β), including standard error (SE) in italics, and significance (p-value) of Relaxometry metrics (MWF, R_1_ and R_2_) vs. Hypertension and age across 14 WM ROIs. The multiple regression model is given by: MRI ∼ β_0_ + β_age_ × age + β_Hypertension_ × Hypertension + β_Smoking_ × Smoking + β_SBP_ × SBP + β_sex_ × sex + β_Diabetes_ × Diabetes + β_Cholesterol_ × Cholesterol, where MRI corresponds to MWF, R_1_ or R_2_. The regression model accounted for sex, smoking status, diabetes status and hypertension as categorical variables. Bolded p-values indicate statistical significance (p < 0.05) or close-to-significance (p < 0.1), after FDR correction.

| ROIs | MWF | | $R_1$ | | $R_2$ | |
| --- | --- | --- | --- | --- | --- | --- |
|  | Hypertension | Age | Hypertension | Age | Hypertension | Age |
| | $\beta$ (SE) $\times 10^{-2}$ | $\beta$ (SE) $\times 10^{-4}$ | $\beta$ (SE) $\times 10^{-5}$ | $\beta$ (SE) $\times 10^{-6}$ | $\beta$ (SE) $\times 10^{-3}$ | $\beta$ (SE) $\times 10^{-5}$ |
| WB | -1.99 (0.64), **0.005** | -6.30 (1.25), <**0.001** | -4.59(1.70), **0.013** | -1.04 (0.33), **0.004** | -1.32 (0.41), **0.004** | -4.51 (0.81), <**0.001** |
| FL | -2.34 (0.65), **0.003** | -7.74 (1.27), <**0.001** | -4.71(1.79), **0.013** | -1.42 (0.35), **0.001** | -1.57 (0.42), **0.002** | -5.26 (0.82), <**0.001** |
| OL | -1.58 (0.81), **0.059** | -4.35 (1.59), **0.008** | -3.17(1.85), **0.096** | -0.69 (0.36), **0.068** | -1.30 (0.51), **0.016** | -3.65 (1.00), <**0.001** |
| PL | -2.21 (0.71), **0.005** | -7.11 (1.40), <**0.001** | -3.95(1.82), **0.038** | -1.31 (0.36), **0.001** | -1.42 (0.49), **0.007** | -5.30 (0.95), <**0.001** |
| TL | -1.97 (0.75), **0.013** | -5.62 (1.46), <**0.001** | -4.35(1.74), **0.018** | -0.97 (0.34), **0.007** | -1.36 (0.48), **0.007** | -4.00 (0.94), <**0.001** |
| CRB | -0.76 (0.68), 0.264 | -3.82 (1.33), **0.006** | -2.75(1.67), 0.102 | -0.43 (0.33), 0.208 | -0.32 (0.46), 0.489 | -3.19 (0.90), **0.001** |
| CC | -2.06 (0.64), **0.005** | -6.56 (1.25), <**0.001** | -7.01(1.73), **0.001** | -1.20 (0.34), **0.001** | -1.76 (0.51), **0.002** | -5.81 (1.00), <**0.001** |
| IC | -1.62 (0.65), **0.019** | -6.68 (1.28), <**0.001** | -5.11(1.65), **0.009** | -0.97 (0.32), **0.005** | -0.90 (0.38), **0.023** | -4.71 (0.74), <**0.001** |
| CR | -2.96 (0.80), **0.003** | -8.51 (1.56), <**0.001** | -6.93(1.87), **0.002** | -1.33 (0.37), **0.001** | -1.74 (0.46), **0.002** | -5.20 (0.91), <**0.001** |
| CP | -1.12 (0.58), **0.059** | -1.92 (1.13), **0.091** | -4.78(1.67), **0.010** | 0.003 (0.33), 0.994 | -0.52 (0.44), 0.251 | -2.60 (0.85), **0.003** |
| TR | -2.18 (0.75), **0.007** | -7.31 (1.47), <**0.001** | -5.10(1.77), **0.010** | -1.26 (0.35), **0.001** | -1.42 (0.47), **0.006** | -4.64 (0.91), <**0.001** |
| FOF | -2.40 (0.71), **0.004** | -7.56 (1.38), <**0.001** | -5.49(1.78), **0.009** | -1.23 (0.35), **0.001** | -1.58 (0.45), **0.002** | -5.55 (0.88), <**0.001** |
| LF | -2.25 (0.73), **0.005** | -6.42 (1.43), <**0.001** | -4.81(1.81), **0.013** | -1.17 (0.36), **0.002** | -1.34 (0.45), **0.007** | -4.08 (0.89), <**0.001** |
| F | -2.16 (0.71), **0.005** | -7.16 (1.40), <**0.001** | -5.13(1.80), **0.010** | -1.20 (0.35), **0.002** | -1.61 (0.46), **0.002** | -5.04 (0.89), <**0.001** |

MWF, myelin water fraction; $R_1$ , Longitudinal rate of relaxation; $R_2$ , Transverse rate of relaxation; WM, white matter; ROI, region-of-interest; SBP, systolic blood pressure; FDR, false discovery rate; WB, whole brain; FL, frontal lobe; OL, occipital lobe; PL, parietal lobe; TL, temporal lobe; CRB, cerebellum; IC, internal capsule; CR, corona radiata; CC, corpus callosum; TR, thalamic radiation; FOF, fronto-occipital fasciculus; LF, longitudinal fasciculus; F, forceps.

**Table 3.**
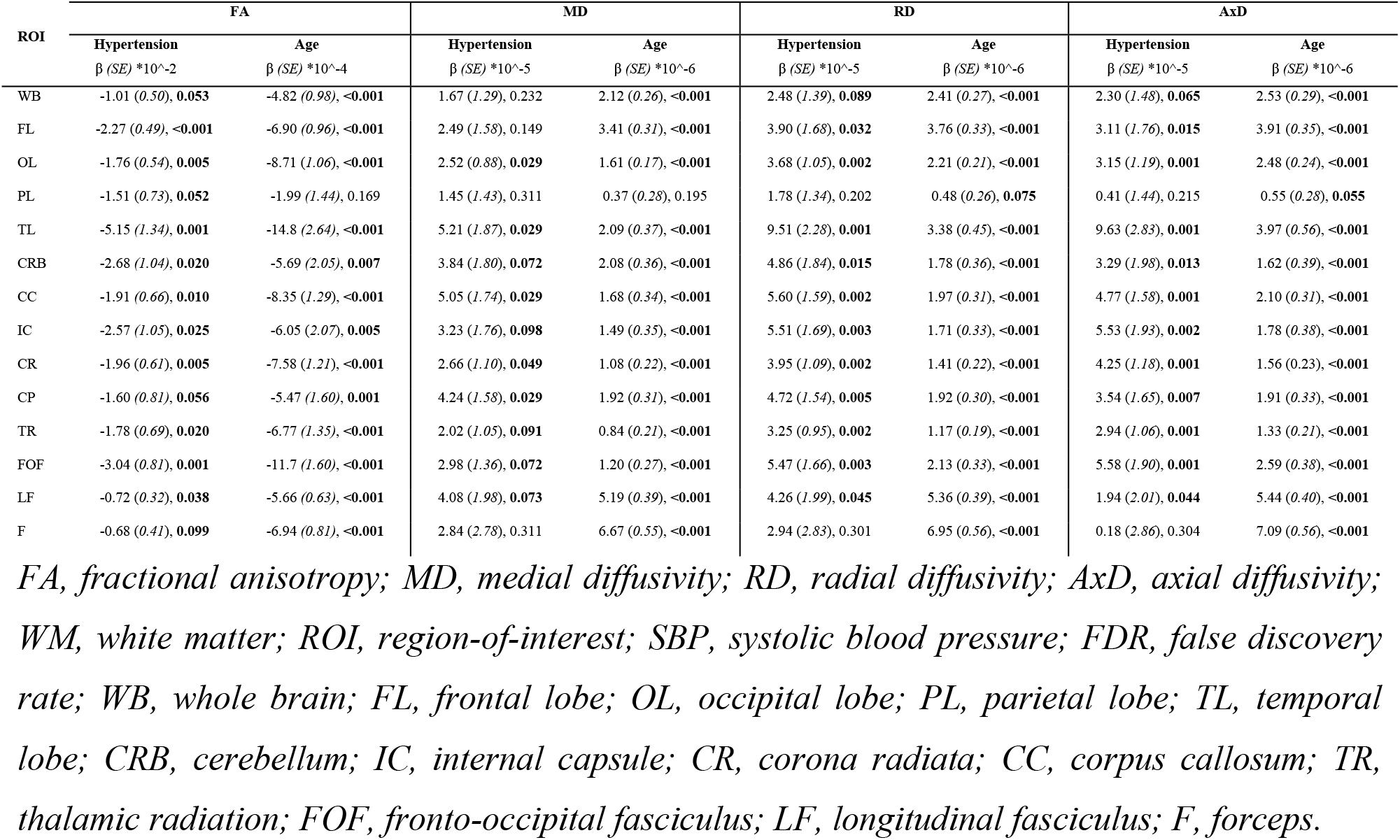
Regression coefficient (β), including standard error (SE) in italics, and significance (p-value) of Diffusion Tensor Imaging metrics (FA, MD, RD and AxD) vs. Hypertension and age across 14 WM ROIs. The multiple regression model is given by: MRI ∼ β_0_ + β_age_ × age + β_Hypertension_ × Hypertension + β_Smoking_ × Smoking + β_SBP_ × SBP + β_sex_ × sex + β_Diabetes_ × Diabetes + β_Cholesterol_ × Cholesterol, where MRI corresponds to FA (Fractional Anisotropy), MD (Medial Diffusivity), RD (Radial Diffusivity) and AxD (Axial Diffusivity). The regression model accounted for sex, smoking status, diabetes status and hypertension as categorical variables. Bolded p-values indicate statistical significance (p < 0.05) or close-to-significance (p < 0.1), after FDR correction.

**Figure 1.**
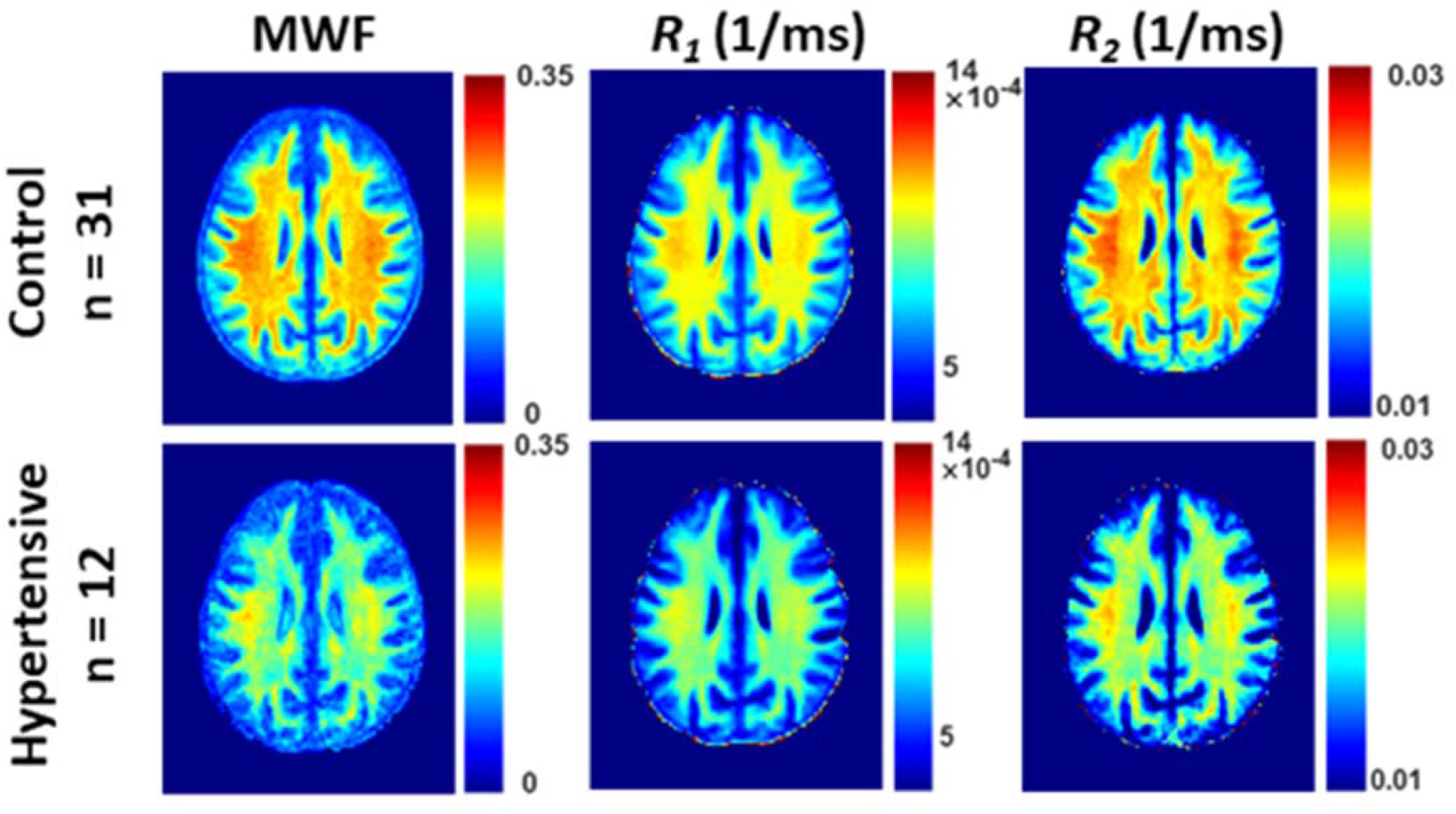
Examples of MWF, R_1_ and R_2_ parameter maps averaged across participants drawn from a limited age range (70-94 yrs.) either hypertensive or non-hypertensive to mitigate the effect of age. Results are shown for a representative slice. Visual inspection indicates that, overall hypertensive patients exhibit lower regional MWF, R_1_ and R_2_ values, as compared to controls. MWF, myelin water fraction; R_1_, longitudinal relaxation rate; R_2_, transverse relaxation rate.

Similarly, Figure 2 shows the FA, MD, RD and AxD DTI parameter maps of either hypertensive or non-hypertensive participants within an age range of 70-94 years. Again, this limited age range is used to minimize the potential effect of age on derived DTI parameter maps for this qualitative analysis. Visual inspection indicates that, overall, hypertensive participants exhibit lower FA and higher MD, RD, and AxD values. In other words, hypertension is associated with higher diffusivities and a lower level of water diffusion. These qualitative results provide further support that hypertension is associated with reduced microstructural white matter integrity.

**Figure 2.**
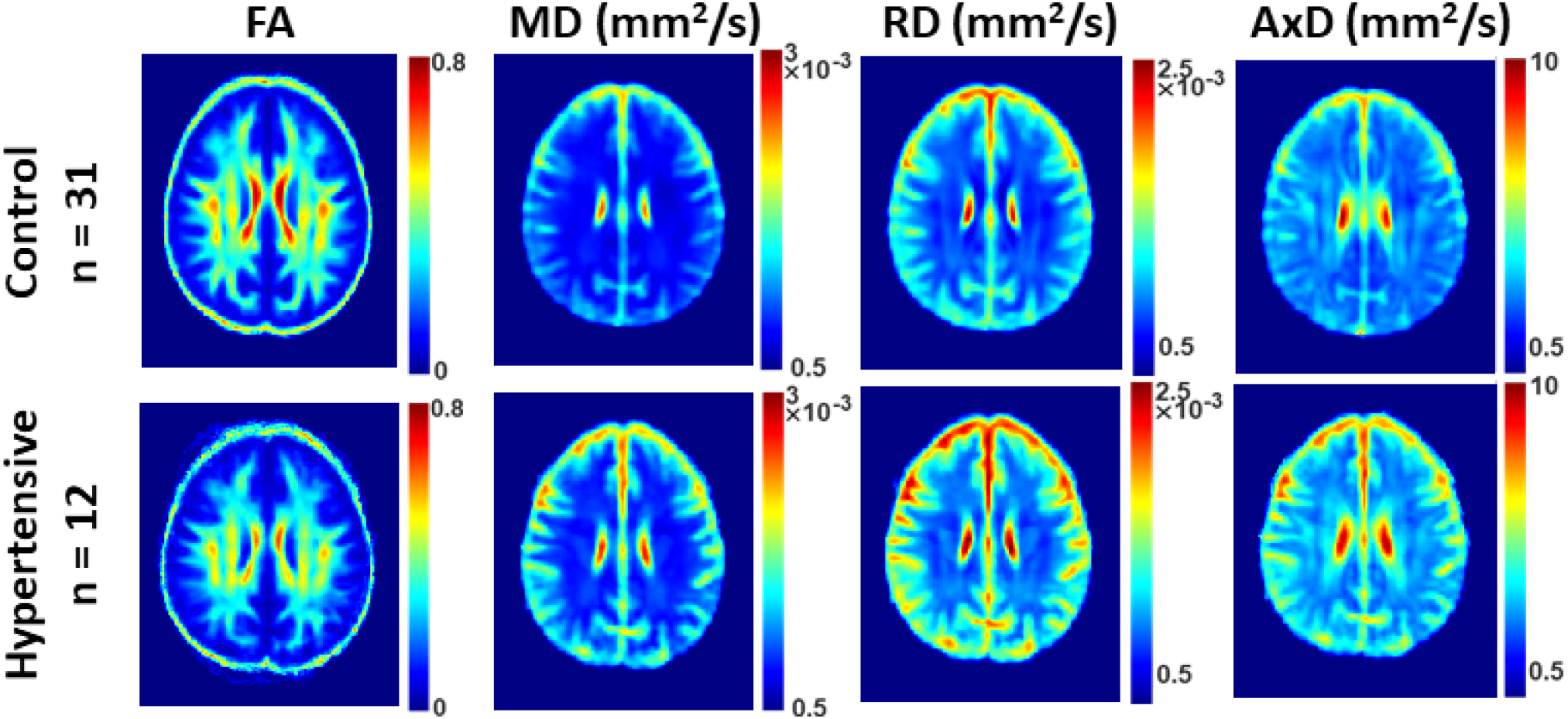
Examples of FA, MD, RD and AxD parameter maps averaged across participants drawn from a limited age range (70-94 yrs.) to mitigate the effect of age. Results are shown for a representative slice. Visual inspection indicates that, overall, hypertensive participants exhibit lower fractional anisotropy (FA) and higher medial diffusivity (MD), radial diffusivity (RD) and axial diffusivity (AxD). FA, fractional anisotropy; MD, medial diffusivity; RD, radial diffusivity; AxD, axial diffusivity.

Table 2 summarizes the results of the multiple regression analysis of MWF, *R*_*1*_ and *R*_*2*_ *vs*. hypertension and age in 14 WM ROIs. In agreement with Figure 1, there are significant (*p* < 0.05), or close-to-significant (*p* < 0.1), negative correlations after FDR correction, between hypertension and MWF or *R*_*1*_ in all regions except for the cerebellum, and negative correlations between hypertension and *R*_*2*_ in all regions but the cerebellum and cerebral peduncles. It was also found that the corpus callosum and the corona radiata exhibited the steepest slopes in the correlations between hypertension and MWF, *R*_*1*_ and *R*_*2*_. Furthermore, as expected, age was found to be a significant covariate with hypertension and exhibited negative slopes with respect to all metrics except for *R*_*1*_ in the cerebral peduncles.

Table 3 summarizes the results of the multiple regression analysis of FA, MD, RD and AxD *vs*. hypertension and age in the 14 WM ROIs studied after controlling for relevant covariates. There are significant (*p* < 0.05), or close-to-significant (*p* < 0.1), negative correlations after FDR correction, between FA and hypertension and positive correlations with MD, RD and AxD in most ROIs investigated. Here, we found that the steepest slopes in the correlation between hypertension and FA, MD, RD and AxD were found in the temporal lobe, fronto-occipital fasciculus, and the internal capsules. We note that in contrast to the results of the multicomponent relaxometry analysis, less ROIs were found to be statistically significant between hypertensive and control groups, with the parietal lobe and forceps regions found to be insignificant in the correlation between hypertension and diffusivity metrics. However, the overall trend of the data follows the paradigm of impaired white matter microstructural integrity. Finally, in all ROIs investigated, the effect of age was found to be significant with respect to all metrics except for FA and MD in the parietal lobe.

## DISCUSSION

In this MRI study, using advanced multicomponent relaxometry and DTI analyses for both direct and indirect measurements of myelin content, we found that the hypertension status is associated with lower regional myelin content as measured by the MWF, *R*_*1*_, *R*_*2*_, and DTI metrics (FA, MD, RD, and AxD). These regional associations were observed in a cohort of well-characterized, cognitively unimpaired, adults. These results provide further evidence of the association between a well-known cardiovascular risk factor, specifically hypertension, and cerebral white matter tissue integrity in the absence of cognitive impairment. Furthermore, to our knowledge, this is the first investigation to indicate a direct association between hypertension and myelin deterioration, as measured by a specific proxy of myelin content (*i*.*e*., MWF). In our analysis, we found that hypertension was associated with higher MD, RD and AxD values and lower MWF, FA, *R*_*1*_ and *R*_*2*_ values. Our DTI results agree with previous DTI studies that have also shown a connection between cardiovascular risk factors, especially hypertension, and decreased cerebral microstructural integrity (9-12, 41-43).

While our relaxometry results, in conjunction with our DTI results, do not prove causality, they support the paradigm that hypertension impairs white matter microstructural integrity, especially the myelination process (5, 44). Indeed, studies have revealed association between increased arterial stiffness and hypertension during the aging process (45-47). One of these paradigms suggests that vascular dysregulation due to potential synergetic effects of arterial remodeling and blood pressure may lead to transient reductions in cerebral blood flow, consequently resulting in transient decreased glucose transport into brain and hypoxia, and concomitant myelin injury (32). Indeed, recent works have demonstrated that deficits in cerebral blood flow are directly linked to reductions in cerebral tissue integrity. This association is present to a greater extent with white matter tissue damage (25, 48, 49). In fact, myelin maintenance through oligodendrocyte metabolism is an energy-demanding process, and therefore myelin homeostasis is particularly sensitive to hypoperfusion and consequent hypoxia or lack of essential nutrients for myelin synthesis (50, 51). Recent *in-vitro* studies have shown that oligodendrocytes are substantially more vulnerable to hypoxia and reduced supply of substrates that provide energy as can occur from hypoperfusion, when compared to other glial cells such as microglia and astroglia (52). Furthermore, it has been shown that hypertension also interferes with perivascular glymphatic drainage and blood–brain permeability. This would result in reduced drainage of toxic metabolites that adversely impact oligodendrocytes, the main cells synthesizing and maintaining myelin in the brain (53). Finally, interruption in the myelination process could result from chronic inflammation. Indeed, animal studies have demonstrated that chronically elevated blood pressure leads to adverse glial activation and increased brain inflammatory mediators that can be harmful to the myelin sheets and the normal functioning of oligodendrocyte cells (54). Nevertheless, despite these potential plausible mechanisms, further studies, especially of a longitudinal nature, are still required to shed light on the association between blood pressure and myelination.

We found the steepest slopes in the correlations between hypertension and MWF, *R*_*1*_ and *R*_*2*_, in the corpus callosum, fronto-occipital fasciculus and corona radiata cerebral regions (Table 2). Numerous studies have found that these brain regions are particularly susceptible to microstructural damages due to elevated blood pressure (55-57). Interestingly, these brain structures have also been shown to exhibit higher sensitivity to the cerebral blood supply (12, 20, 58, 59). For example, the corpus callosum has an especially high level of metabolic demand, receiving blood from the anterior communicating, anterior pericallosal, and posterior cerebral arteries (60). Although ischemia in this region is rare due to the trifurcated nature of the vascular pathway, the energy demanding process of myelination could be impeded from even minor changes in blood flow, such as those that occur from hypertension (61, 62).

Longitudinal studies have found that anti-hypertensive medication has a protective effect on the brain and helps to reduce the rate of cognitive decline and neurodegeneration, including in Alzheimer’s disease (63, 64). These studies consistently find that elevated blood pressure in midlife is more closely associated with cognitive decline when compared with elevated blood pressure in late life (65, 66). This could possibly be due to the slow progression of hypertensive arterial remodeling eventually leading to reduced blood flow postischemia (5). Interestingly, among the 27 hypertensive subjects in our study, 23 subjects were taking antihypertensive medication at the time of the scan. Although it is unclear whether the anti-hypertensive medication had some level of a protective effect on these participants, it is interesting to note that participants undergoing treatment still had significantly lower myelin content or higher microstructural damage in many of the regions analyzed (Table 1). We conjecture that this may be due to either microstructural damage being done prior to the treatment of the anti-hypertensive medication or as a demonstration of the possible limitations of the antihypertensive medication on protecting the overall cerebral microstructure long term (Figure 1,2).

Although our investigation examined a relatively large cohort and used advanced MRI methodology to probe myelin content and obtain diffusion metrics, our work has certain limitations. The cross-sectional nature of the study precludes us from drawing any causal link between hypertension and demyelination; future longitudinal studies are needed to further support this potential association. We also note that the causal relationship between hypertension and myelination is difficult to determine as hypertension commonly occurs concomitant with many other cardiovascular risk factors, and we cannot control for all of them in our limited multiple linear regression model given the cohort size. Finally, determination of MR parameters can be biased due to several biological and methodological factors. These include, but are not limited to, the effects of magnetization transfer between macromolecules and free water protons, exchange between water pools, J-coupling, off-resonance, spin locking effects, water diffusion within different compartments, flow, temperature, hydration, internal gradients, and architectural features, including fiber fanning or crossing (48).

## CONCLUSION

This study provides new insights into the association between hypertension and axonal demyelination among cognitively normal individuals spanning a wide age range. This work motivates further investigations to elucidate the extent to which hypertension and myelination are related in the pathological progression of neurodegenerative diseases, including in Alzheimer’s disease and dementias.

## Data Availability

All data and codes are available through a direct request from the corresponding author.

## ACKNOWLEDGEMENT

This research was supported entirely by the Intramural research Program of the NIH, National institute on Aging.

## Notes

### Competing Interest Statement

The authors have declared no competing interest.

### Author Declarations

Protocol was approved by the MedStar Research Institute and the National Institutes of Health Intramural Ethics committees, and all examinations were performed in compliance with the standards established by the National Institutes of Health Institutional Review Board.

